# Using Joint Longitudinal and Time-to-Event Models to Improve the Parameterization of Chronic Disease Microsimulation Models: an Application to Cardiovascular Disease

**DOI:** 10.1101/2024.10.27.24316240

**Authors:** John Giardina, Sebastien Haneuse, Ankur Pandya

## Abstract

**Background:** Chronic disease microsimulation models often simulate disease incidence as a function of risk factors that evolve over time (e.g., blood pressure increasing with age) in order to facilitate decision analyses of different disease screening and prevention strategies. Existing models typically rely on incidence rates estimated with standard survival analysis techniques (e.g., proportional hazards from baseline data) that are not designed to be continually updated each model cycle. We introduce the use of joint longitudinal and time-to-event to parameterize microsimulations to avoid potential issues from using these existing methods. These joint models include random effects regressions to estimate the risk factor trajectories and a survival model to predict disease risk based on those estimated trajectories. In a case study on cardiovascular disease (CVD), we compare the validity of microsimulation models parameterized with this joint model approach to those parameterized with the standard approaches.

**Methods:** A CVD microsimulation model was constructed that modeled the trajectory of seven CVD risk factors/predictors as a function of age (smoking, diabetes, systolic blood pressure, antihypertensive medication use, total cholesterol, HDL, and statin use) and predicted yearly CVD incidence as a function of these predictors, plus age, sex, and race. We parameterized the model using data from the Atherosclerosis Risk in Communities study (ARIC). The risk of CVD in the microsimulation was parameterized with three approaches: (1) joint longitudinal and time-to-event model, (2) proportional hazards model estimated using baseline data, and (3) proportional hazards model estimated using time-varying data. We accounted for non-CVD mortality across all the parameterization approaches. We simulated risk factor trajectories and CVD incidence from age 70y to 85y for an external test set comprised of individuals from the Multi-Ethnic Study of Atherosclerosis (MESA). We compared the simulated to observed incidence using both average survival curves and the E50 and E90 calibration metrics (the median and 90th percentile absolute difference between observed and predicted incidence) to measure the validity of each parameterization approach.

**Results:** The average CVD survival curve estimated by the microsimulation model parameterized with the joint model approach matched the observed curve from the test set relatively closely. The other parameterization methods generally performed worse, especially the proportional hazards model estimated using baseline data. Similar results were observed for the calibration metrics, with the joint model performing particularly well on the E90 metric compared to the other models.

**Conclusions:** Using a joint longitudinal and time-to-event model to parameterize a CVD simulation model produced incidence predictions that more accurately reflected observed data than a model parameterized with standard approaches. This parameterization approach could lead to more reliable microsimulation models, especially for models that evaluate policies which depend on tracking dynamic risk factors over time. Beyond this single case study, more work is needed to identify the specific circumstances where the joint model approach will outperform existing methods.

## 1 Introduction

Simulation models are often used to conduct health decision analyses when there is no single dataset that contains all the relevant information to compare the treatment and policy options.^1^ Simulations can be especially useful for analyses of chronic illnesses like cardiovascular disease (CVD), where there are a large number of potential treatment and prevention strategies (e.g., at what age and risk level should cholesterol screening be started) and the effects of these options need to be compared over the lifetime of an individual.^2^ No single clinical trial will be able to provide all the information needed to identify the best strategy, so simulation models can be used to integrate all available data sources, including both observational and trial data, to help decision-makers choose the best option.^1^

In order for these models to be useful when making decisions, however, they need to accurately represent the disease process. For example, if a model is going to evaluate at what age low-density lipoprotein (LDL) cholesterol screening should be started to guide statin initiation, it needs to accurately predict how LDL levels evolve as individuals age and how those levels relate to cardiovascular disease (CVD) risk (which in turn informs the potential effectiveness of statin treatment).^3^ This study analyzes potential approaches to developing chronic disease microsimulation models so that the predicted progression of disease incidence and risk factors over time matches that actual natural history of the disease under current treatment choices as closely as possible. Existing models often use previously published risk scores or standard risk prediction techniques to estimate disease incidence over time, but these approaches are not necessarily appropriate to track the continual, dynamic changes in risk factors and disease risk included in a microsimulation model. We introduce the use of joint longitudinal and time-to-event models to parameterize microsimulation models; these joint models simultaneously estimate the progression of risk factors over time and their relationship to disease incidence, so they are particularly well suited to parameterize chronic disease microsimulation models. We assess how well this approach compares to other available parameterization methods in a case study of CVD microsimulation model.

The remainder of this paper is structured as follows. First, background on the parameterization of chronic disease microsimulation models is reviewed. Second, joint longitudinal and time-to-event models are introduced as a way to parameterize simulation models, and a simple example comparing this approach to existing parameterization methods is discussed. Third, we implement the joint model and other parameterization approaches in a CVD case study and estimate the validity of the microsimulation models developed under these different approaches. Finally, we review the results of the case study and the implications for the development of future chronic disease microsimulation models.

## 2 Background

For this study, we focus on the parameterization of microsimulation models that have the general structure shown in Figure 1. In these models, a hypothetical cohort of individuals are given a set of baseline risk factors (e.g., blood pressure, smoking status); each model cycle, these risk factors are updated as the individuals age and the updated risk factors are used to predict the risk of a disease event (e.g., stroke, heart attack) within the cycle.

**Figure 1:**
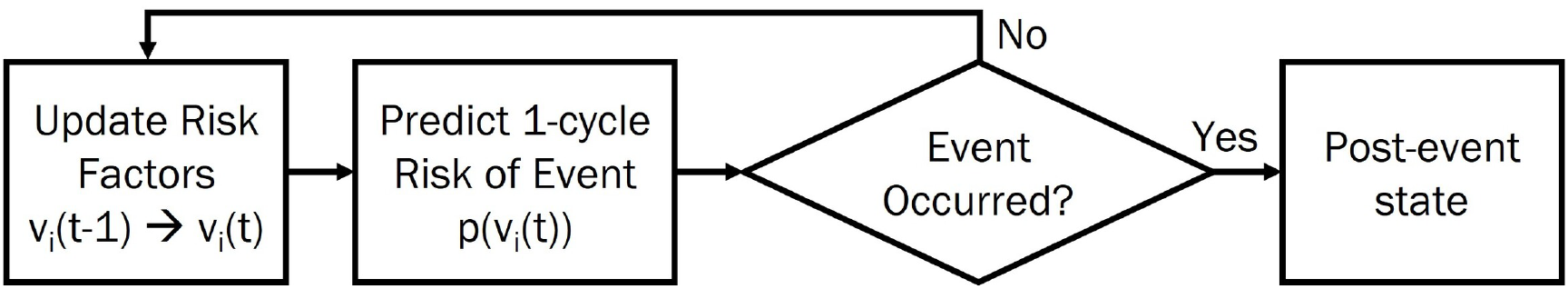
Stylized microsimulation model structure for chronic disease prevention and screening models *v*_*i*_(*t*): Vector of risk factors values at time *t* (e.g., blood pressure, smoking status) for individual *i* in the model population. *p*(*v*_*i*_(*t*)): Probability of an event (e.g., stroke, heart attack) from time *t* to *t* + 1 based on risk factor values

This type of model structure is needed when a microsimulation model is going to be used to evaluate treatment and prevention policies that are based on risk factor values. For example, a model might be used to evaluate when and how often we should screen people for atrial fibrillation (AF) in order to initiate anticoagulant treatment for stroke prevention. To evaluate these policies, we need to know what the prevalence of AF is at each time point and how that prevalence is related to future stroke risk.

There are two key parameters needed to build models with the structure in Figure 1: (1) a function to update risk factor values from one model cycle to the next, and (2) a function that calculates the probability of a disease event based on those risk factor values. The ideal dataset to calculate these parameters would be individual-level data from a cohort study, with risk factor measurements repeated at least as frequently as the length of each model cycle. The function to update risk factor values over time could be estimated with either standard regression techniques for longitudinal data (e.g., fixed or random effects regression) or by directly using the risk factor trajectories of individuals from the data within the model (potentially with matching or reweighting to fit desired model population). Similarly, the probability of the disease event in each cycle could be calculated by regressing each set of risk factor measurements against the occurrence of the disease event within the timeframe of one cycle. Some models with access to regularly collected longitudinal data take this approach. For example, the RAPIDS model for diabetes progression was parameterized using electronic medical records and claims data from the US Department of Veterans Affairs.^4^ Biomarker and disease data were generally available on the same interval as the model cycle and any missing data was interpolated, so lagged variable regression models were used to estimate the progression of risk factors and disease events.

Unfortunately, often the only data available to parameterize simulation models are from cohort studies that have limited collection of risk factor values (e.g., once per year over a five-year period) relative to the cycle length and time horizon of the microsimulation model (e.g., one year cycle length over a 30-year time horizon). These datasets usually include longer-term collection of time-to-event data for the disease event, but there can be a large gap between the last collected risk factor measurements and the time that the event occurs. This means that, in order to parameterize the relationship between risk factor values and the event risk within each model cycle, assumptions need to be made to fill in the gap between risk factor measurements and the time of the disease event.

Existing models that have adapted the structure shown in Figure 1 have often done this by estimating the risk of a disease event as a function of risk factors using survival analysis techniques like proportional hazards models, and then separately estimating how those risk factors evolve over time by regressing risk factor values against age (e.g., the CVD PREDICT and UKPDS Outcomes diabetes models^5,6^). Standard survival analysis techniques, however, usually implicitly assume that the risk factor values used to estimate the survival model stay constant between measurements.^7^ This can create issues when implementing these risk prediction functions within microsimulation models that continually update disease risk from new, updated risk factor values (discussed further in the Illustrative Example section below). Joint longitudinal and time-to-event models, which have previously been developed in the biostatistics literature, have the potential to overcome these issues by combining regressions of risk factor values over time with the estimation of a survival model.^7^ The approach, however, has never been previously used to parameterize a microsimulation model.

A joint model has two components: (1) a random effects regression to predict risk factor values over time, and (2) a survival model that incorporates those risk factor predictions to estimate the risk of a disease event at a given time. Given the correct specification assumption (e.g., linear relationship between age and risk factor value), the regressions can more accurately interpolate and extrapolate the progression of these risk factors values compared to an assumption that the values stay constant after an observation. In a sense, the use of regressions to “fill-in” the gaps between risk factor measurements is a way to approximate the ideal dataset for microsimulation parameterization discussed above, where there are risk factor measurements available for every model cycle. Some microsimulation models have used similar approaches to extrapolate risk factor values to the event time. For example, the CEPAC HIV model parameterized the rate of opportunistic infections (OI) among individuals with HIV as a function of CD4 cell count by (1) extrapolating the last observed CD4 cell count using the age fixed effects coefficient from a mixed effects regression, and then (2) directly estimating the OI rate in each model cycle based on that extrapolation.^8^ Full joint longitudinal and time-to-event models, however, avoid this two-step process. If there is longitudinal data available and the assumptions underlying the model are correct (e.g., normal distribution for the random effects, correct functional specifications), joint models have the capacity to account for risk factor trends within patients, correlations between multiple risk factors, and measurement error.^7^

## 3 Illustrative Example

In this section, we provide an illustrative example of how joint longitudinal and time-to-event models have the potential to improve on existing methods for the prediction of disease risk within a chronic disease microsimulation model. We compare the joint model approach to a standard proportional hazards model approach that either (1) only uses risk factor values collected at baseline in a cohort study (“baseline model”) or (2) uses repeated risk factor measurements, but treats each measurement as a different observation that is not connected to the value of the previous measurement (“time-dependent model”).^7^ The baseline model is similar to the Framingham risk score used to parameterize the CVD PREDICT model, which has been used to evaluate CVD prevention strategies like statin initiation.^6,9^ The Framingham risk score only used baseline risk factor measures from the Framingham cohorts to estimate an accelerated failure time model.^10^ The time-dependent model is similar to the diabetes complication risk functions used to parameterize the UKPDS Outcomes model, which was developed to predict outcomes for individuals with type 2 diabetes mellitus.^5^ Those risk functions were developed with a proportional hazards model that included time-varying covariates but, based on the model description, did not adapt the risk estimation process to account for potential changes in risk factor values between or after observation times.^5^

For this example, assume that there is available cohort data that includes time-to-event data for incident disease events and irregularly collected longitudinal risk factor data that includes measurement error, represented by the dots in Figure 2. The goal is to use this data to parameterize a microsimulation model similar to Figure 1, so we need to correctly predict the relationship between risk factor levels and the hazard of a disease event. This will allow the microsimulation to continually update the risk of a disease event based on the progression of risk factors over time. In this example, the true underlying trajectory of the risk factor values is a linear increase over time (Figure 2, dashed line), and there is a positive relationship between the risk factor value and the event hazard (Figure 3, dashed line).

**Figure 2:**
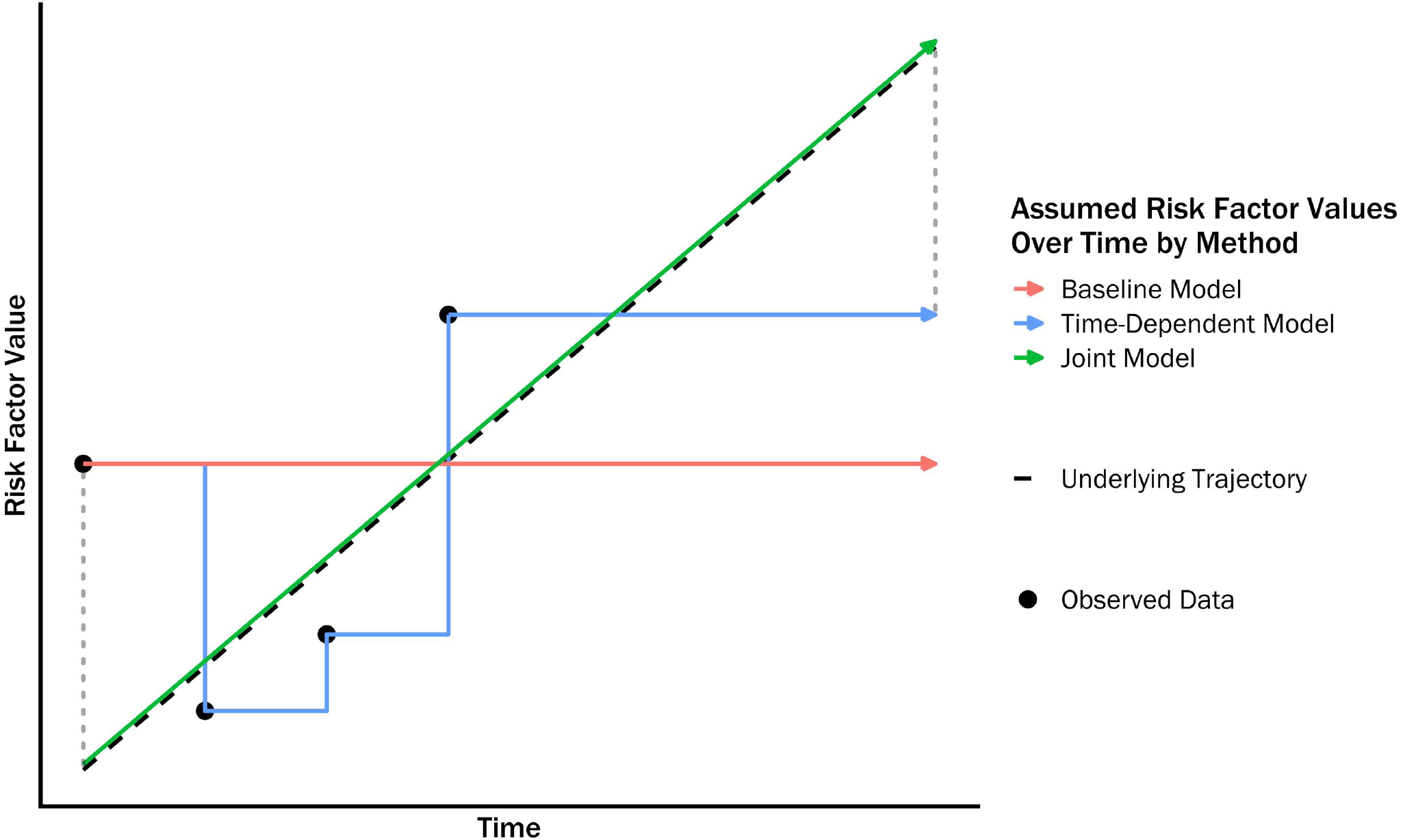
Assumptions about risk factor values over time by risk estimation method from illustrative example Underlying trajectory (dashed line): The actual risk factor values increase linearly over time but there is measurement error, as shown by the observed data points. Baseline Model (red line): Assumes starting risk factor value stays constant over time, and that values are measured without error. Time-Dependent Model (blue line): Assumes last-observed risk factor value is constant until next observation, and that values are measured without error. Joint Model (green line): Fits random effects regression model to interpolate extrapolate risk factor trajectory, and accounts for random measurement error.

**Figure 3:**
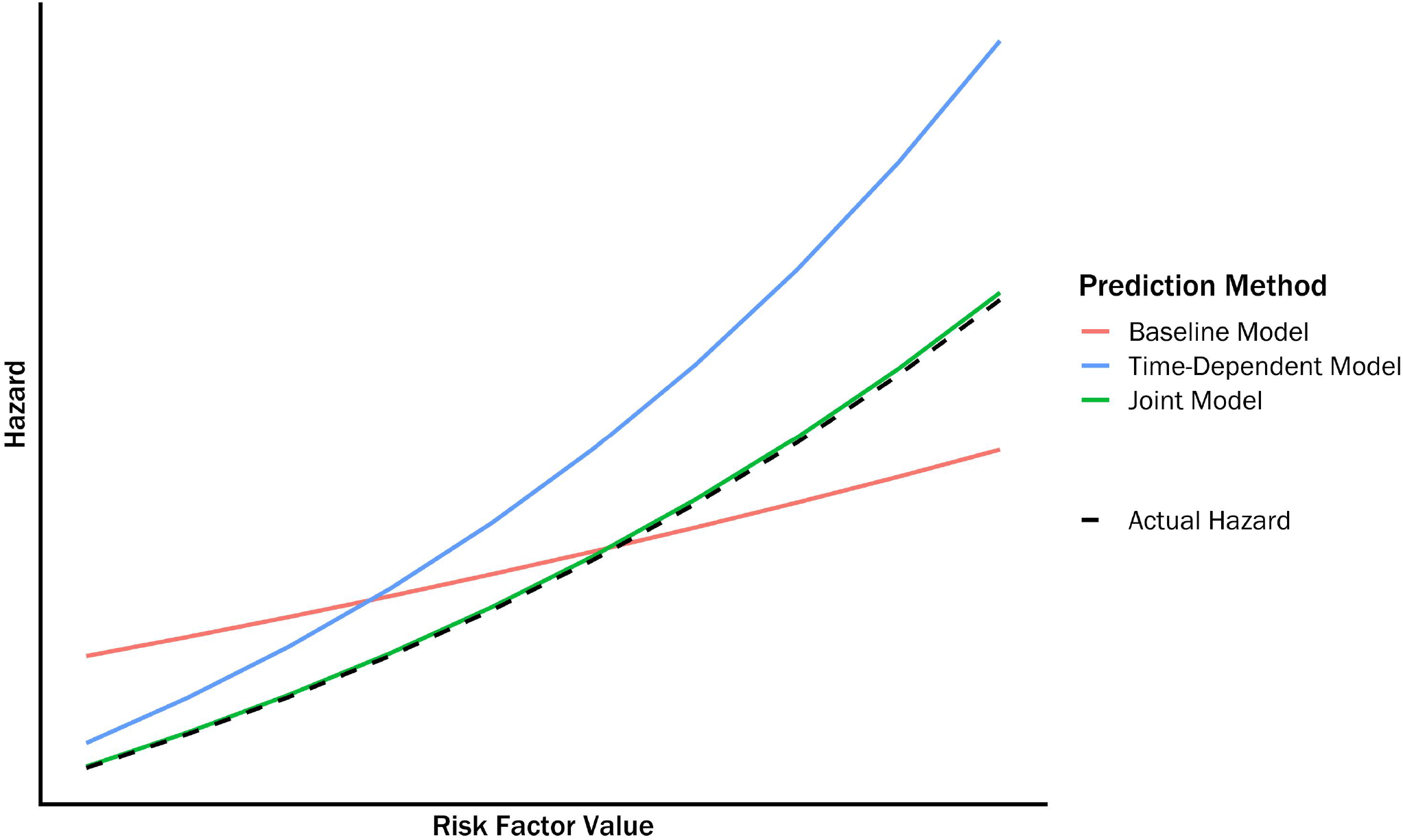
Estimated hazard by risk estimation method from illustrative example Actual Hazard (dashed line): Increases along with increases in risk factor values. Baseline Model (red line): Attributes future increases in hazard to an underlying time-trend because only lower initial risk factors are used to estimate risk, so the model underestimates the relationship between the risk factor values and event risk. Time-Dependent Model (blue line): Since risk factor value observations stop midway through follow-up, attributes future increases in hazard in second-half of follow-up to lower carried-forward risk factor values. This overestimates the relationship between the risk factor values and event risk. Joint Model (green line): Attributes future increases in hazard to extrapolated increased risk factor values, so the model accurately estimates relationship between risk factor and event risk.

The baseline model only observes the first risk factor value, so it essentially assumes that value stays constant over the whole follow-up period (Figure 2, red line). The time-dependent model does not account for connections between the longitudinal measurements (e.g., time trend), so it assumes that the values are constant between observation times and that the last measured value stays constant for the remainder of the follow-up period (Figure 2, blue line). The joint model, on the other hand, fits a random effects regression to the observed values (Figure 2, green line). If the specification of the regression is correct, it can capture the true underlying trajectory of the risk factor values, interpolating and extrapolating the risk factor values over time and accounting for any random measurement error.

For the baseline and time-dependent models, these assumptions lead to incorrect estimates of the relationship between the risk factor value and event hazard. The baseline model attributes future increases in hazard to an underlying time-trend unrelated to the risk factor because it only uses the lower, initial risk factors to estimate the hazard. This means that the baseline model underestimates the relationship between the risk factor values and event risk (Figure 3, red line). For the time-dependent model, it only observes risk factor values through the first half of follow-up; this means that it attributes future increases in hazard in the second half to lower carried-forward risk factor values. This leads to an overestimate of the relationship between the risk factor values and event risk (Figure 3, blue line). Since the joint model, however, makes correct assumptions about the underlying trajectory of the risk factor, it attributes future increases in hazard to extrapolated increased risk factor values and accurately estimates the relationship between the risk factor and event risk (Figure 3, green line).

## 4 Case Study

While a joint modeling approach clearly has the potential to provide benefits over standard methods in a simple, constructed context, it is important to assess whether those benefits still apply when parameterizing a microsimulation model with actual data. To do this, we built a basic microsimulation model of CVD and parameterize it using a joint longitudinal and time-to-event model, along with the existing baseline and time-dependent survival model approaches discussed in the preceding section. We then validate the model using observed CVD incidence data to assess which parameterization approaches best replicate the natural history of CVD.

### 4.1 Methods

#### 4.1.1 Cardiovascular Disease Microsimulation Model

Following the stylized model structure shown in Figure 1, we constructed a basic microsimulation model for the natural history of primary CVD incidence with a 1-year cycle length that estimated (1) the progression of CVD risk factors and predictors over time; and (2) the yearly incidence of CVD as a function of those risk factors. We included nine risk factors and predictors in the model, based on the covariates included in the Pooled Cohort Equations (PCE): age, sex, race (African American vs. non-African American), current smoking status, diabetes mellitus, systolic blood pressure, current use of antihypertensive medication, total cholesterol, HDL, and statin use.^11^

We constructed three versions of the model for each of the three different parameterization approaches (i.e., joint model, baseline model, time-dependent model). For all of the models, we estimated the risk factor trajectories and CVD incidence over 15 years with a starting cohort of 70-year-old individuals, and included non-CVD mortality risk as a competing risk for CVD (the structure for the model version including non-CVD mortality risk is shown in Figure S.1 in the Supplementary Materials).

#### 4.1.2 Data

We parameterized and evaluated the model using data from two cohort studies obtained from the BioLINCC repository at the National Heart, Lung, and Blood Institute: the Atherosclerosis Risk in Communities (ARIC) study and the Multi-Ethnic Study of Atherosclerosis (MESA).^12,13^ This data includes time-to-event data for incident CVD and death, and longitudinal data collected at repeated clinic visits, including measurements of the risk factors listed in the previous section (i.e., current smoking status, diabetes mellitus, systolic blood pressure, current use of antihypertensive medication, total cholesterol, HDL, and statin use). We defined incident CVD as the first occurcence of myocardial infarction, stroke, or fatal coronary heart disease; we do not include angina or heart failure in our definition of CVD, reflecting the defition of “hard” atherosclerotic cardiovascular disease (ASCVD) events used in the PCE.^11^ Additional information on these data is included in the Supplementary Materials S.

The data from ARIC was used to parameterize the microsimulation models (*training set*), and the data from MESA was used to validate the model (*test set*). Only individuals who did not have a history of CVD or heart failure at baseline, had non-missing values for the the time to CVD and death data, and had at least one non-missing value for each of the risk factors were used in the training and test sets. For the training set, 9.75% of the individuals in the original ARIC data were removed because of prevalent CVD or heart failure at baseline and 4.51% were removed for other reasons. For the test set, 0.07% of individuals in the original MESA data were removed because of prevalent CVD or heart failure at baseline and 0.44% were removed for other reasons (the MESA study exclusion criteria included baseline CVD, but some individuals were identified after enrollment). Any risk factor information from visits after incident CVD was excluded from the pooled data. The Harvard T.H. Chan School of Public Health Institutional Review Board determined that use of the ARIC and MESA data was not human subjects research (IRB18-1797).

The final training set included 12,865 individuals, and risk factor data was repeatedly collected for these patients across a total of 53,163 visits. A total of 2,872 incident CVD events (22% of individuals) were observed in this data, with each individual being observed for an average of 22.32 years (SD: 8.86) before incident CVD, death, or censoring. Additional descriptive statistics for the training set are provided in Table S.1 in the Supplementary Materials S.

Since the test set was used to validate the microsimulation model, only individuals in MESA who had risk factor observations prior to age 70 and who were alive and had no prevalent CVD at age 70 were allocated to the test set. This allowed us to populate a baseline cohort for the microsimulation model with a starting age of 70 that matched the test set. The final test set included 2,659 individuals, with 192 obvserved incident CVD events. Individuals were observed for an average of 6.80 years (SD: 4.14) before incident CVD, death, or censoring. Additional information on the construction of the test set is included in the Appendix, and descriptive statistics are provided in Table S.2 in the Supplementary Materials S.

#### 4.1.3 Parameterization Approaches

The key parameters within the model are (1) the functions to estimate the values of the risk factors in each 1-year model cycle, and (2) and the probability of incident CVD within that year based on those risk factors. The functions to estimate the risk factor values were derived with two methods: (1) “stand-alone” regression functions, derived directly from the available longitudinal data, similar to methods used to parameterize existing models;^6,14^ and (2) the random effects regressions from a joint longitudinal and time-to-event model. For both of these methods, the value of the seven risk factors (i.e., smoking status, diabetes, systolic blood pressure, use of antihypertensive medication, total cholesterol, HDL, and use of statins), were estimated as a function of age, sex, and race (African American vs. non-African American) with a linear specification. The stand-alone regression functions were estimated for each risk factor in isolation (without considering correlations between risk factor values), and only included random intercepts; this approach is similar to the one taken by Leal et al. [14] for the UKPDS Outcomes Model (version 2). The random effects regressions from the joint longitudinal-survival model also included random intercepts and slopes, and a covariance matrix to account for correlations between the random effects of different risk factors. The stand-alone regression functions were estimated using the lme4 R package,^15^ and the random effects regression from the joint model were estimated using the JMbayes2 R package.^16^ Additional information on the estimation and implementation of the risk factor updating functions is included in the Supplementary Materials S.

The functions to estimate the probability of CVD within each cycle were derived using three methods: (1) the survival component from a joint longitudinal-survival model (joint model); (2) a proportional hazards model using baseline data from the training set (baseline model); and (3) a proportional hazards model using all longitudinal data from the training set (time-dependent model). The joint, baseline, and time-dependent risk models were all estimated on the age time scale (rather than time from baseline) and included cubic splines for age to account for the varying impact of age on CVD risk. For all these approaches, we accounted for non-CVD mortality as a competing risk using the cause-specific hazards approach. The joint model was estimated using the JMbayes2 R package,^16^ and the baseline and time-dependent models were estimated using the survPen R package.^17^ Additional information on the estimation of the CVD risk functions is included in the Supplementary Materials S.

#### 4.1.4 Starting model Population

The starting population for the model was constructed to match the individuals in the test set sot the CVD incidence in that set could be used to validate the model. The baseline risk factor values for each individual (i.e., the risk factor values at age 70) were estimated by using the risk factor updating functions to extrapolate the risk factor values observed in the test set prior the starting age. When the random effects regressions from the joint model were used as the risk factor updating functions, this extrapolation process included updating the random effects distributions for each individual so that any risk factor trends observed prior to the starting age persisted throughout the model runs (through the updated random slope component). Additional information on how the risk factor updating functions were used to parameterize the starting population is included in the Supplementary Materials S.

#### 4.1.5 Model Implementation

We ran 3 versions of the model for each of the primary methods of estimating CVD probability: the joint model, baseline model, and time-dependent model. The joint model version used the risk factor updating functions already estimated as part of the joint model, and the baseline and time-dependent model versions used the standalone regression functions as the risk factor updating functions.

The primary output from these versions of the model was the estimated cumulative incidence of CVD for each individual in the model population for every year from the starting to ending age. Note that, since the microsimulation model structure was relatively straightforward, we calculated the estimated cumulative incidence directly from the estimated risk prediction functions rather than implementing a full microsimulation that discretely simulated CVD incidence (i.e., where an individual ends a model run with CVD or no CVD). This reduced the computation time needed to run the model, and by construction calculates what the estimated cumulative CVD incidence would be from a full microsimulation with discrete events; more details are provided in the Supplementary Materials S.

#### 4.1.6 Model Validation Comparisons

Two approaches were used to validate the predictions of cumulative CVD incidence from each version of the model. First, the cumulative incidence was averaged across all the individuals in the model population to construct an average CVD survival curve; this average predicted survival curve was compared to the observed survival curve from the test set to assess whether each model predicted population-level CVD incidence correctly. The observed survival curve was estimated non-parametrically using the Nelson-Aalen estimator for cumulative hazard, with the log-log approach used to calculate a 95% confidence interval, and accounted for non-CVD mortality using the cause-specific hazards approach.

Second, the median and 90th percentile absolute differences between the observed and model-predicted CVD risk across all individuals in the test set were calculated at a range of age landmarks (from 1 years after the starting age until age 85, at 1-year increments). These absolute differences reflect the E50 and E90 calibration metrics described by Austin et al. [18], and are a measure for how closely the predicted CVD incidence matches observed CVD incidence across the range of incidence levels in the population. While we cannot directly observe the CVD incidence rate or risk for each individual (we only observe the occurrence of CVD or no CVD over the set time frame), a proxy for the “observed” CVD incidence for each individual is calculated by fitting a proportional hazards model to a cubic spline for the complementary log-log transformation of the predicted incidence; the observed incidence for each individual is the incidence estimated from this fitted model.^18^ This proportional hazards model is fit using the Fine-Gray subdistribution hazard approach to account for non-CVD mortality, following the recommendation from Austin et al. [19]. In general, this approach to calculating calibration metrics can be thought of as a smoothed version of other metrics that compare predicted to observed incidence across discrete bins.^18^ All the model code and validation analyses were run with the R programing language, version 4.4.1.^20^

### 4.2 Results

#### 4.2.1 CVD Survival Curves

Overall, the estimated average CVD survival curve from the joint model microsimulation was closer to the observed test set curve than those estimated from microsimulations parameterized with the baseline and time-dependent approaches (Figure 4); the predicted CVD survival curve from the joint model consistently fell within the 95% confidence interval of the observed survival curve. Between the two other methods, the time-dependent model generally performed better than the baseline model, but models overpredicted CVD incidence compared to the observed incidence in the test cohort.

**Figure 4:**
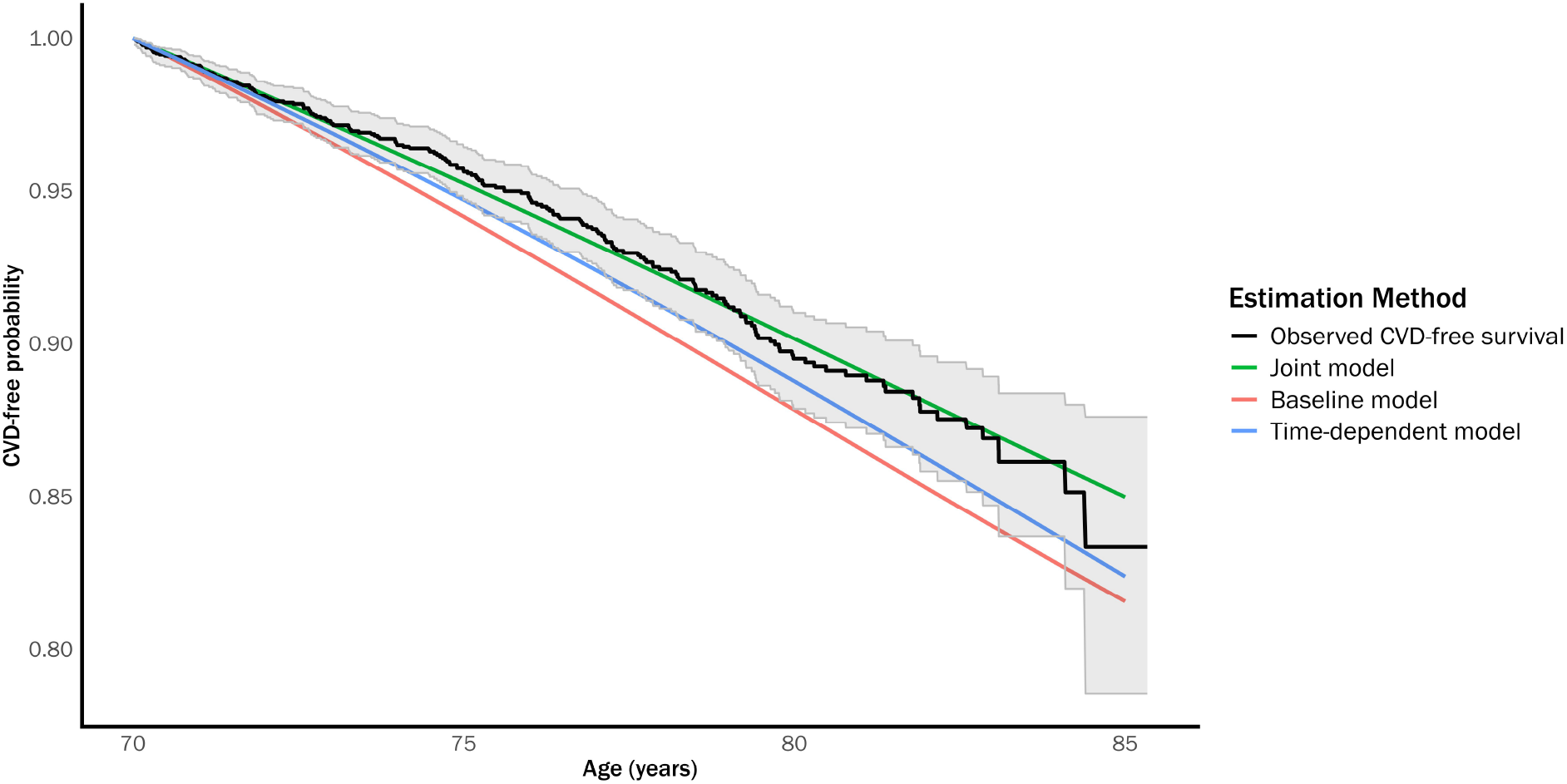
Average CVD survival curves for microsimulation models with age 70 starting cohort Gray region is 95% confidence interval of observed CVD survival curve calculated with the log-log approach.

##### Calibration Metrics

Generally, the median absolute differences between estimated and observed CVD risk from the joint model microsimulation were on par with the same metric from the time-dependent model simulations, and outperformed the baseline simulation (Figure 5). The joint model microsimulation performed noticeably better than both the time-dependent and baseline versions when comparing the 90th percentile absolute differences, indicating that the time-dependent and baseline model versions made more extreme “outlier” predictions that deviated from the observed risk compared to the joint model.

**Figure 5:**
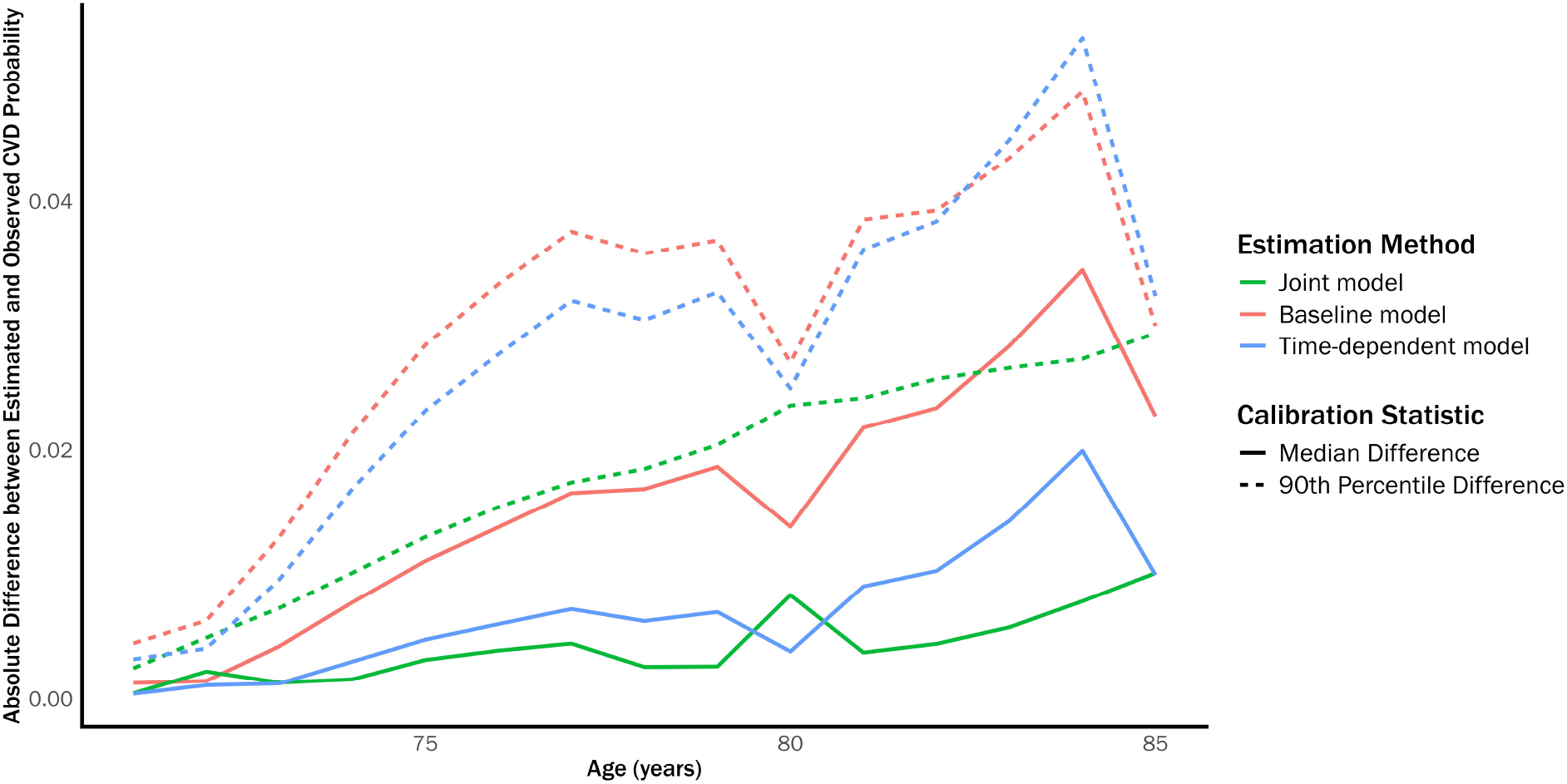
Calibration metrics for microsimulation models with age 70 starting cohort

## 5 Discussion

This study assessed methods to parameterize chronic disease microsimulation models that track both the progression of risk factors and the evolution of disease risk over time; we introduced joint longitudinal and time-to-event models as an approach to parameterize this type of model and compared it to existing methods. In a case study of a CVD microsimulation model, we found the joint model approach to parameterization accurately replicated the observed CVD survival curve in an external test set (Figure 4). It also significantly outperformed microsimulations parameterized with standard survival models that only used baseline data from a cohort study (baseline model), and performed better than or on par with standard models that used longitudinal data from these studies (time-dependent model), depending on the comparison metric used.

Parameterizing microsimulation models with joint longitudinal and time-to-event models has the capacity to provide more valid and accurate simulations compared to existing approaches, which could in turn lead to more accurate decision analyses that are conducted using these simulations. The time-dependent modeling approach slightly outperformed the joint model on the E50 calibration metric at a few age landmarks for some of the model versions (i.e., Figure 5), but in general the joint model approach better predicted CVD incidence both on average and across individual risk levels.

There are limitations to this study. First, this analysis only evaluated the use of joint models for microsimulation parameterization in one case study. This means more work is needed to identify under what specific conditions joint models perform better than existing parameterization approaches. In order to guide future model development, it would be helpful to know what the relationship between risk factor progression, disease incidence, and the structure of the available data needs to be for a joint model approach to be useful; this will likely require additional applied and simulated analyses. Additionally, we only evaluated joint models in comparison to a specific set of existing approaches (i.e., baseline and time-dependent survival models, with stand-alone regressions for risk factor updating). There are a number of variations to these existing approaches (e.g., Markov model approach that does not link disease risk with risk factor updating),^21^ and future work comparing these variations to the joint model approach could also help identify when using a joint model will most improve model validity.

Relatedly, it is not clear from this single case study when the assumptions underlying the joint model approach (e.g., functional specification of risk factor trajectories, normal distribution of random effects) would impede performance of the model. Similarly, it is unclear if the cause-specific hazards approach to accounting for competing risks will always produce accurate predictions. Lau et al. [22] suggests that the Fine-Gray sub-distribution hazards approach may result in better predictions, but this option was not available in the software package we used to estimate the joint model. Future work could explore the impact of these assumptions and modeling choices on the validity of microsimulation models parameterized with the joint model approach.

Finally, there are limitations to implementing joint models. For instance, it can be computationally intensive to fit and draw from the posterior distributions of the model. This should be taken into account when integrating the joint modeling approach into a microsimulation model, which are often computationally intensive on their own. Additionally, fitting a joint model requires longitudinal data with repeated observations across individuals. This type of dataset is not always available when constructing a microsimulation model, although this may change if cohort study data (such as the studies included in the BioLINCC repository), claims data, and electronic health records become more widely available for use in research (although the biases inherent in these data sources would also need to be evaluated through the model development process).

In addition to these limitations, there are also a number of other potential directions for future research into how joint longitudinal and time-to-event models can best be utilized within decision analytic models. First, because the joint modeling approach includes regressions for the risk factor trajectories, dynamic strategies that account for patient histories can be integrated into decision models parameterized with joint models.^7^ For example, the risk of CVD could be predicted using the area under the systolic blood pressure trajectory curve to capture accumulated risk over time; this relationship could be used within the simulation model to assess screening strategies that take past blood pressure readings into account, rather than just the measurement at a single point in time. Next, although it was not discussed in this study, joint models are usually estimated within a Bayesian framework. Another potential direction for future work is to investigate how to best integrate the posterior distributions estimated from a joint model with other Bayesian approaches commonly used in decision modeling, like value of information analyses or Bayesian calibration techniques.^23,24^

## 6 Conclusion

This study introduced the use of joint longitudinal and time-to-event models to parameterize chronic disease microsimulations models, and compared the validity of microsimulations developed with this joint model approach to other existing methods in a case study on CVD. The microsimulation models parameterized with a joint model generally showed increased validity compared to existing methods, indicating that this approach could be used to conduct more accurate decision analyses of chronic disease screening and prevention strategies.

## Data Availability

All data used in the present study are available from the Biologic Specimen and Data Repository Information Coordinating Center (BioLINCC) at the National Heart, Lung, and Blood Institute (NHLBI).

https://biolincc.nhlbi.nih.gov/studies/aric/

https://biolincc.nhlbi.nih.gov/studies/mesa/

## Manuscript Information

### Author Contributions

*Concept and design:* All authors.

*Acquisition, analysis, or interpretation of data:* All authors.

*Drafting of the manuscript:* Giardina.

*Critical revision of the manuscript for important intellectual content:* All authors.

*Statistical analysis:* Giardina.

*Obtained funding:* Pandya.

*Administrative, technical, or material support:* Giardina.

*Supervision:* Pandya.

### Funding and Support

The authors were supported by the National Institutes of Health (grant No. R01NS104143 to Pandya) and Harvard University (dissertation completition fellowship to Giardina).

The funders had no role in the design and conduct of the study; collection, management, analysis, and interpretation of the data; and preparation, review, or approval of the manuscript. The content of this study is solely the responsibility of the authors and does not necessarily represent the official views of the NIH, Harvard University, or the ARIC, CHS, MESA, and Framingham Offspring investigators.

## Acknowledgments

This work was prepared using ARIC, CHS, MESA, and Framingham Offspring Research Materials obtained from the NHLBI Biologic Specimen and Data Repository Information Coordinating Center.

## S Supplementary Materials

### S.1 Included Studies

The ARIC study recruited individuals 45-64 years-old across four communities in the US; the available ARIC data included seven clinic visit cycles (the first four conducted between 1987-1998, the fifth, sixth, and seventh conducted 2011-2019).^1,2^ The MESA study recruited individuals 45-84 years old who were free of cardiovascular disease at baseline across six communities in the US; the available MESA data included five clinic visit cycles conducted between 2000-2011.^3,4^ The data collection procedures and other details of these studies have been previously published.^1,3^

### S.2 Test Set Construction

As noted in the main text, the construction of the test set depended on the starting age of the cohort for the microsimulation model it would be used to validate. To evaluate models that had a cohort starting at age 70, the test set only included individuals who had risk factor information available prior to age 70 (so baseline risk factors at age 70 could be estimated) and did not have incident CVD, death, or censoring until after age 70.

### S.3 Data Notation

In order to describe the approaches we took to parameterize the microsimulation model in this study, we use the following notation for the data available from the pooled cohort studies: (1) repeated observations for each of the seven risk factors (i.e., current smoking status, diabetes status, systolic blood pressure, use of antihypertensive medication, total cholesterol, HDL, and use of statins) for each individual are represented by *y*_*ikq*_ for the *k* observation of risk factor *q* for individual *i*; (2) the ages of the individuals at each observation are represented by *age*_*ik*_, and the sex and race (African American vs. non-African American) are represented by *sex*_*i*_ and *race*_*i*_, respectively; (3) the age when we start observing each individual for CVD is represented by *T*_*i,start*_; and (4) the age when an individual experiences incident CVD, dies, or is censored is represented by *T*_*i,stop*_, along with an indicator for whether they had CVD, died, or were censored, represented by *δ*_*i*_ (equals 0 if censored, 1 if incident CVD was observed, and 2 if death was observed).

### S.4 Joint Longitudinal-Survival Model

The joint longitudinal-survival model (joint model) estimated in this study was comprised of two components: (1) a random effects regression component to estimate the evolution of risk factors over time, and (2) a survival component to estimate the hazard of CVD based on those risk factors. All components of the joint models used in this study were estimated with the JMbayes2 package using the default priors.^5^ We summarize the specification and assumptions for the model below, relying heavily on Rizopoulos [6] for notation and the specifics of the joint model approach.

For each of the seven risk factors included in the model, the random effects regression had the following linear specification:

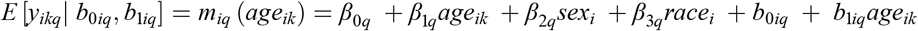

Here, 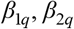 and 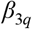 are the fixed effects of age, sex, and race, respectively, on the risk factor *q*, and *b*_0*iq*_ and *b*_1*iq*_ are the random intercept and slope, respectively, of risk factor *q* for individual *i*. Each risk factor is assumed to be normally distributed, conditional on the random effects: 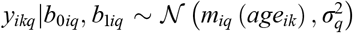.Note that we made this assumption even for the binary risk factors (e.g., smoking status) since other distributional assumptions caused convergence issues for the model. This means that the regressions for these binary variables should be interpreted as linear probability models. Additionally, the random effects across all risk factors are assumed have a multivariate normal distribution with mean 0 and a covariance matrix *D*, which captures the relationships between the different risk factors: **b**_*i*_ = (*b*_0*i*1_, *b*_1*i*1_, …, *b*_0*i*7_, *b*_1*i*7_) ∼ 𝒩 (0, *D*).

The survival component had the following specification:

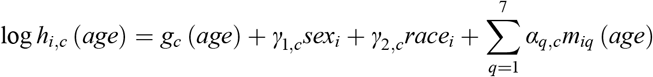

Here, *h*_*i,c*_ be the hazard function for cause *c* and individual *i* as a function of *age*, where *c* = 1 for CVD and *c* = 2 for death, *g*_*c*_ (*age*) are cubic splines of age with ten interior knots, *γ*_1,*c*_ and *γ*_1,*c*_ are the regression coefficients relating sex and race to the log hazard for cause *c, m*_*iq*_ (*age*) are the mean risk factor values for individual *i* and risk factor *q* at the given age from the random effects regression, and *α*_*q,c*_ are regression coefficients relating the risk factor values to the log hazard for cause *c*. Note that the JMbayes2 package used to estimate this model estimates the splines for age as penalized B-splines.^5^

Within the joint model framework, it is assumed that the following distributions are independent conditional on the random effects (**b**_**i**_) and all other parameters (**θ**): the distributions of the different risk factors (i.e.,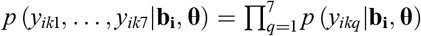; the distribution of the risk factors across different times (i.e., *p* (*y*_*i*1*q*_, *y*_*i*2*q*_, …| **b**_**i**_, **θ**) = Π_*k*_ *p* (*y*_*ikq*_|**b**_**i**_, **θ**)); and the distribution of the risk factors and CVD event times (i.e., *p* (*y*_*ikq*_, *T*_*i,stop*_, *δ*_*i*_|**b**_**i**_, **θ**) = *p* (*y*_*ikq*_|**b**_**i**_, **θ**) *p* (*T*_*i,stop*_, *δ*_*i*_|**b**_**i**_, **θ**)).^6^

Given these assumptions and taking a Bayesian approach to the model estimation, the posterior for the model estimated by the JMbayes2 package is:^7^

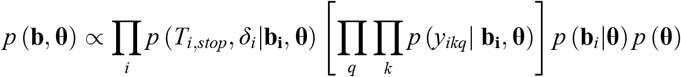

Here, **b** is the vector of all the random effects **b**_**i**_, the distribution of the risk factors and random effects are provided by the normal distribution assumptions described above, and the distribution of the event times and causes can be expressed as a function of the cause-specific hazards:

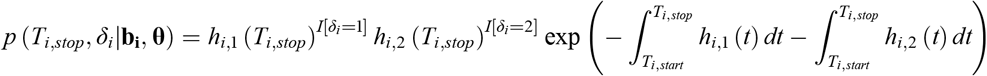

The exp 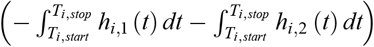 term captures the likelihood that individual *i* survives from *T*_*i,start*_ to *T*_*i,stop*_ without CVD or death, and the 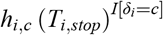 terms capture the likelihood that individual *i* will experience an event with cause *c* exactly at time *T*_*i,stop*_ if we observe an event at that time (i.e., *δ*_*i*_ = *c*).

### S.5 Baseline Model

The baseline model estimated in this study was a parametric survival model fit using the survPen package,^8^ which implemented methods described by Fauvernier et al. [9]. The model used the following specification for the log-hazard function:

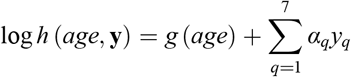

Here, *g* (*age*) are penalized restricted cubic splines of age with ten interior knots, *y*_*q*_ is the value of the *q* risk factor, and *α*_*q*_ are regression coefficients relating the risk factor values to the log hazard. Note that, unlike the joint model described above, there is no explicit link in this specification between the age and the time when the risk factor values were recorded. In order to implement this hazard function in the microsimulation model, as described in the Model Implementation section below, it needs to be assumed the risk factor value is measured at the given age. To estimate the baseline model, however, we only use the baseline level risk factors for each individual (similar to standard survival analysis approaches), which essentially assumes that the baseline risk factor values stay constant across every age. The likelihood used to fit the baseline model is then the following function, where **θ** is a vector of the parameters for the cubic splines for age and the regression coefficients *α*_*q*_, and *y*_*i*1_ is a vector of the baseline risk factor values for individual *i*:

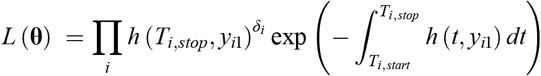

Note that, as implemented by the survPen package, this likelihood function was penalized to smooth the cubic spline terms when fitting the model;^8^ Fauvernier et al. [9] can be referenced for more details. Additionally, the competing risk of death was modeled by taking a cause-specific hazards approach and fitting a separate hazard function for non-CVD mortality. Details of how these two hazard functions were integrated into the microsimulation model are discussed in the Model Implementation section below.

### S.6 Time-Dependent Model

The procedure to estimate the time-dependent model was almost identical to the baseline model, except all the risk factor measurements were included to fit the model, rather than just the baseline measurements. This was done by treating the time from one risk factor observation time to the next as separate time-to-event processes. Assume that individual *i* has *k* = 1, …, *K*_*i*_ risk factor observations; the time at each observation is represented by *T*_*i,k*_, so that *T*_*i,k*_ = *age*_*ik*_ for *k* = 1, …, *K*_*i*_, and the final CVD, death, or censor time is represented by 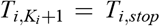 the censoring indicator *δ*_*ik*_ equals 1 if an individual has incident CVD (or non-CVD mortality, for the mortality-specific hazard model) at time *T*_*i,k*_ and equals 0 otherwise. Additionally note that, by construction, *T*_*i*,1_ = *age*_*i*1_ = *T*_*i,start*_. Using the same hazard specification as the baseline model, the following likelihood function is used to fit the model, where *y*_*ik*_ is the vector of the risk factor values for individual *i* taken at observation *k*:

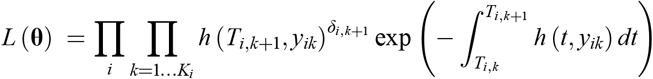

Again, since this model was implemented using the survPen package, the likelihood function was penalized when fitting the model to smooth the cubic spline terms.^8^ The competing risk framework is implemented in the same way as for the baseline model (i.e., a separate hazard function was fit for non-CVD mortality).

### S.7 Parameterizing Risk Factor Updating Functions

Two approaches were used to parameterize the risk factor updating functions in the microsimulation model. The first approach was to fit “stand-alone” regressions to estimate how risk factors progress with age, which is common in existing models.^10,11^ To do this, the following regression was fit for each risk factor separately:

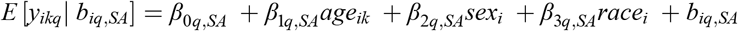

Here, *β*_1*q,SA*_, *β*_2*q,SA*_, and *β*_1*q,SA*_ are the fixed effects of age, sex, and race (African American vs. non-African American) on the risk factor *q* and *b*_*iq,SA*_ is the random intercept for individual *i* (*SA* is added to the subscripts for “stand-alone” to differentiate these parameters from the joint model parameters). Each risk factor is assumed to have a normal distribution, conditional on the random effects:

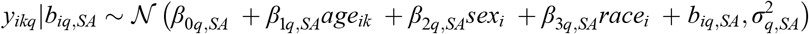

Additionally, note that, to match the assumptions we made for the joint model, the assumption of a normal distribution assumption was made even for the binary risk factors (e.g., smoking status). The random intercepts are assumed to have a normal distribution with mean 0, but we did not model any correlation between the risk factors (as was done in the joint model). To implement these updating functions in the microsimulation model, the age fixed effects were extracted and applied to the baseline risk factor values in the population for the simulation; this is discussed further in the Model Implementation section below.

The second approach to estimate the risk factor updating functions was to use the risk factor functions *m*_*iq*_ (*age*) from the joint model to estimate the progression of risk factors as individuals age. Note that these functions depend on the distribution of the random effects **b**_**i**_, so when this updating function was implemented in the microsimulation model, the distribution of **b**_**i**_ was updated to reflect any information about previous or baseline risk factor values; this is discussed further in the Model Implementation section below.

### S.8 Microsimulation Model Implementation

In order evaluate the different microsimulation model parameterization methods described in this study, we implemented a version of the model structure detailed in Figure S.1 for incident CVD. So that we can evaluate the validity of the model parameterization, we use the test sets as the baseline population for the model; for each individual in the test set, the goal is to estimate the cumulative incidence of CVD at every modeled age 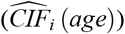,so that we can then compare that to the actual, observed incidence. In the sections below, we discuss how we parameterize the two major components of the model (the risk factor updating functions and CVD risk in each model cycle). Note that we do not implement a full stochastic microsimulation, where an individual is run through the model repeatedly, assigned “CVD” or “no CVD” during each model run, and those counts are averaged to obtain 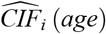.Instead, in order to save computation time, we calculate 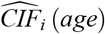 directly using the estimated hazard functions; for the joint model this means we integrate over continuous time, and for the other model versions we multiply the per-cycle probability of CVD. The details of this approach are discussed below.

#### S.8.1 Risk Factor Updating

For each version of the microsimulation model, we estimated the progression of risk factor values for each individual in the simulation from age 70 to age 85. The baseline population for the microsimulation models was drawn from the test set that would be used to validate the model. For each individual *i* in the population, we constructed risk factor updating functions *ŷ*_*iq*_ (*age*) that estimated the value of risk factor *q* at any age after the starting cohort age.

For model versions that used the “stand-alone” regressions to parameterize the risk factor updating function (i.e., the ones that used the baseline and time-dependent CVD survival models), the estimated progression of the risk factor values were calculated by taking the last observed risk factor values for each individual in the test set before the starting age (*y*_*i,q,last*_; e.g., last observation before age 70), and then applying the estimated age fixed effects 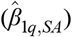 from the stand-alone regressions to extrapolate that value to estimate the risk factor value at the each age in the microsimulation model, where:

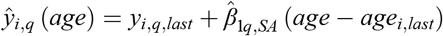

Here, *age*_*i,last*_ is the age of individual *i* at the last observation before the starting age. For example, if the last observed blood pressure value was 125 mmHg at age 65 and the fixed effect of age was an increase of 0.5 mmHg each year, the estimated value at age 70 would be 127.5 mmHg.

For microsimulation version that used the joint model to parameterize the survival model and risk factor updating functions, all the observations prior to the start age were used to update the distribution of the random effects and calculate the expected progression of the risk factors based on that new distribution. That is, if **y**_*i,prior*_ represents the vector of all risk factor measurements for individual *i* in the test set prior to the starting age of the cohort and *p* (**θ**|*data*_*training*_) is the posterior distribution of all the other parameters obtained by fitting the joint model with the training data, then we estimate the risk factor updating functions for individual *i* with:^6^

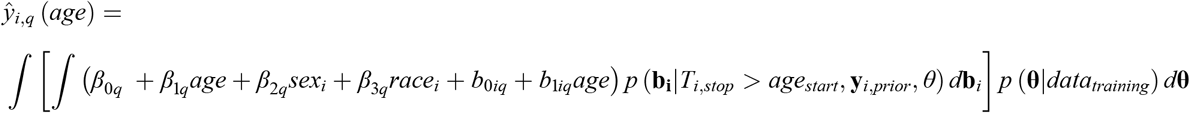

Here, note that *β*_0*q*_, *β*_1*q*_, *β*_2*q*_, and *β*_3*q*_ are components of **θ**, and *age*_*start*_ is the starting age of the cohort for the microsimulation model (i.e., age 70). Conditioning on *T*_*i,stop*_ *> age*_*start*_ is to account for the fact that population used in the model has not had incident CVD or died by the starting age. These predictions were obtained using the predict function in the JMbayes2 R package.^5^

#### S.8.2 Modeling CVD Incidence

After constructing the risk factor updating functions, as described above, we modeled the incidence of CVD for each individual in the test set from the ages 70 to 85, where 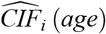 represents the estimated cumulative incidence function for individual *i* (conditional on being CVD-free at the starting age of the cohort).

For versions of the microsimulation model parameterized with the joint model, we calculated this incidence directly from the posterior distribution of the fitted joint model. This means that we are not actually implementing a full microsimulation approach, where individuals are run repeatedly through a stochastic process. Instead, this process estimates the mean incidence we would observe in that full microsimulation but with less computational burden. To estimate the cumulative incidence for each individual, we again update the distribution of the random effects based on the risk factor values observed in the test set prior to the starting age, as was done with the risk factor updating function:^12^

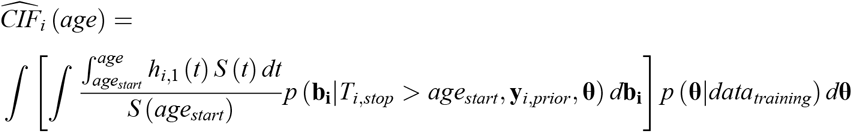

As in the initial formulation of the joint model, 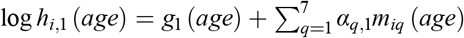 where 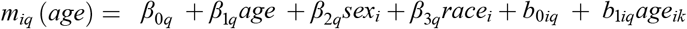. Additionally, 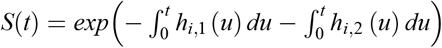 is the overall survival function for both CVD and death, accounting for both cause-specific hazards. Again, note that all the parameters in these functions, aside from *b*_0*iq*_ and *b*_1*iq*_, are components of **θ**. The predictions of the cumulative incidence of CVD were obtained using the predict function in the JMbayes2 R package.^5^

For the versions of the microsimulation model parameterized with the baseline and time-dependent models, let the vector of simulated risk factor progressions of risk factor for individual *i* be *ŷ*_*i*_ (*age*), with the elements *ŷ*_*i,q*_ (*age*) for risk factor *q* from the stand-alone risk factor regressions discussed above. The estimated cumulative incidence of CVD for each individual is calculated with:

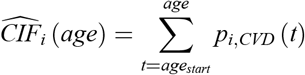

Here, *p*_*i,CVD*_ (*t*) is the overall probability of CVD for individual *i* in the time between *t* and *t* + 1, not conditional on surviving up to that period. This is calculated by multiplying the probability of surviving up until time *t* by the probability of incident CVD between *t* and *t* + 1, conditional on survival until time *t*:

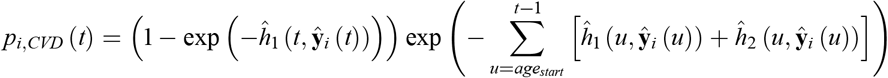

Here, *ĥ*_1_ and *ĥ*_2_ are the cause-specific hazard functions for CVD and non-CVD death estimated by the baseline and time-dependent methods (discussed above), which are a function of age and the risk factor values. The cumulative incidence function for these model versions are estimated discretely, instead of integrating over the relevant time periods, to reflect a microsimulation model that that has a one-year cycle length.

### S.9 Microsimulation Model Validation

#### S.9.1 Average estimated Survival Curve

We obtained the average estimated CVD-specific survival curves reported in the Results section with the following formula (assuming *N* individuals in the test set):

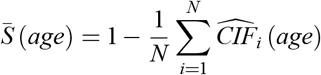

#### S.9.2 Calibration Metrics

The E50 and E90 calibration metrics reported in the Results section were calculated using the methodology from Austin et al. [13]. To calculate these metrics at a given age, we first fit the following proportional hazards model to the observed CVD incidence in the test set, where *n* is a cubic spline function with three degrees of freedom and using a Fine-Gray subdistribution model to account for the competing risk of death:

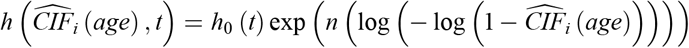

For each individual *i*, we then estimated the “observed” cumulative CVD incidence at the given age by plugging in 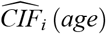 to the fitted model and using the Breslow estimator for the baseline cumulative hazard. With this observed incidence 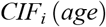,we calculated the absolute difference between the observed and estimated incidence for each individual 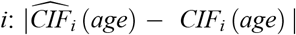.The median of this absolute difference across all individuals is the E50 metric, and the 90th percentile is the E90 metric.

### S.10 Supplementary Figures

**Figure S.1:**
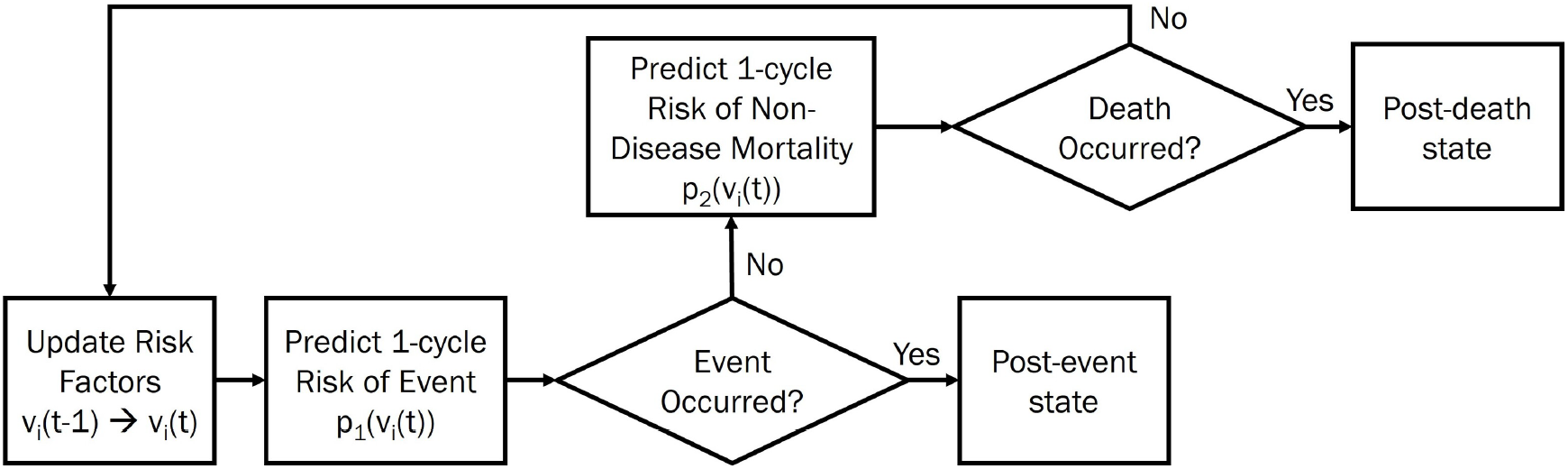
Stylized microsimulation model structure for chronic disease prevention and screening models including non-disease mortality *v*_*i*_(*t*): Vector of risk factors values at time *t* (e.g., blood pressure, smoking status) for individual *i* in the model population. *p*_1_(*v*_*i*_(*t*)): Probability of an event (e.g., stroke, heart attack) from time *t* to *t* + 1 based on risk factor values *p*_2_(*v*_*i*_(*t*)): Probability of mortality not caused by the disease event from time *t* to *t* + 1 based on risk factor values

### S.11 Supplementary Tables

**Table S.1:**
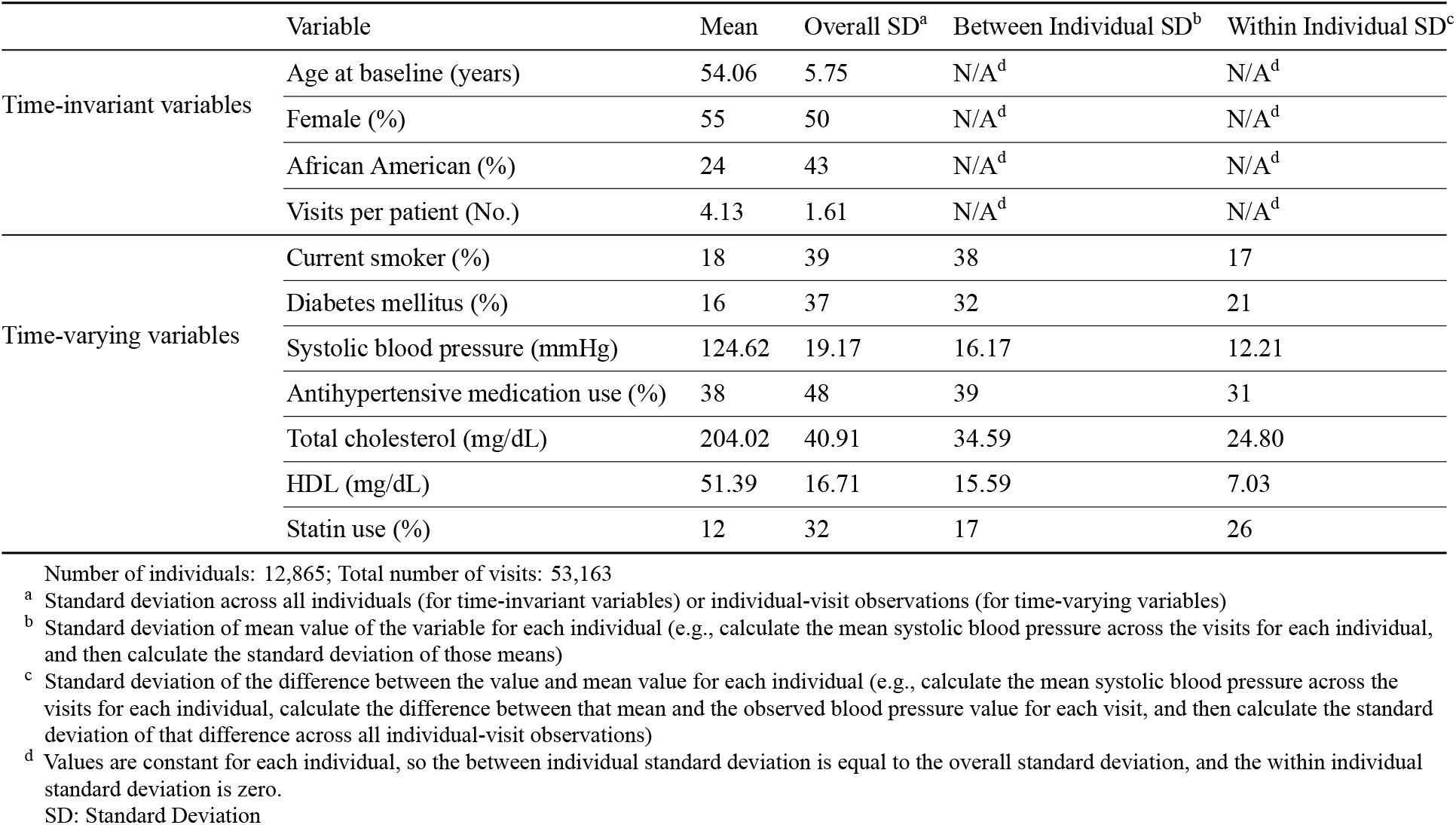
Descriptive Statistics for Training Set

**Table S.2:**
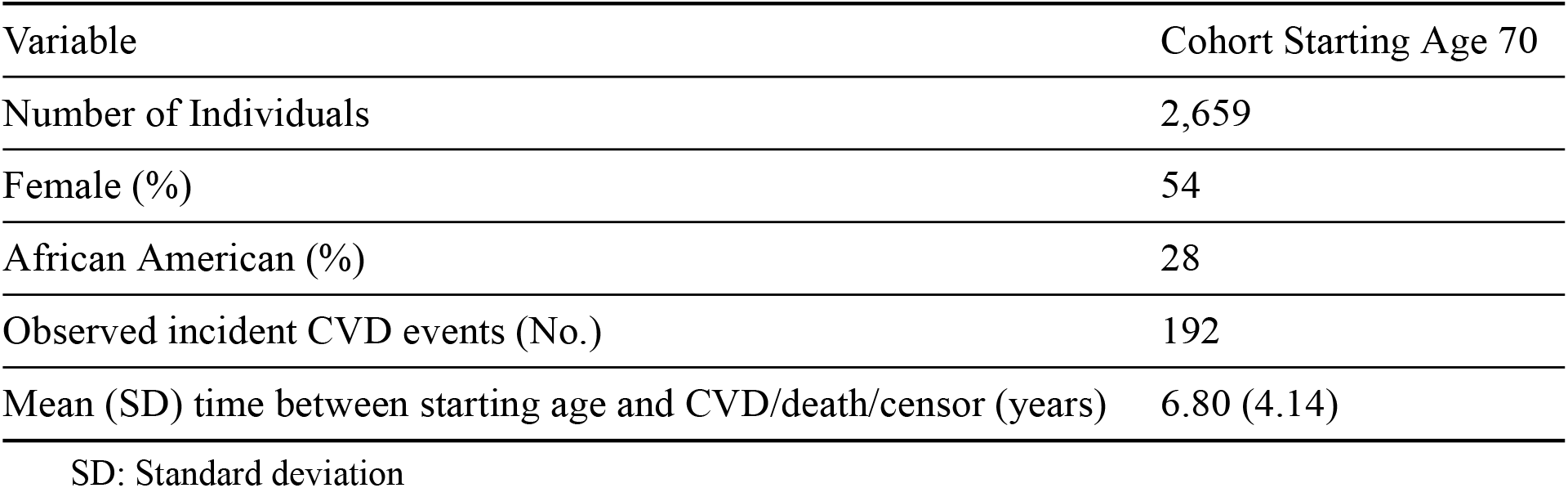
Descriptive Statistics for Test Set

## Notes

### Competing Interest Statement

The authors have declared no competing interest.

### Author Declarations

The Harvard T.H. Chan School of Public Health Institutional Review Board determined that use of the ARIC and MESA data was not human subjects research (IRB18-1797).

## References

1. Kuntz KM, Russell LB, Owens DK, Sanders GD, Trikalinos TA, and Salomon JA. Decision Models in Cost-Effectiveness Analysis. In: Cost-Effectiveness in Health and Medicine. Ed. by Neumann PJ, Ganiats TG, Russell LB, Sanders GD, and Siegel JE. Oxford University Press, 2016. DOI: 10.1093/acprof:oso/9780190492939.003.0005.

2. Unal B, Capewell S, and Critchley JA. Coronary heart disease policy models: a systematic review. BMC Public Health 2006;6:213. DOI: 10.1186/1471-2458-6-213.

3. Eddy DM, Hollingworth W, Caro JJ, et al. Model transparency and validation: a report of the ISPOR-SMDM Modeling Good Research Practices Task Force-7. Med Decis Making 2012;32:733–43. DOI: 10.1177/0272989X12454579.

4. Basu A, Sohn MW, Bartle B, Chan KCG, Cooper JM, and Huang E. Development and Validation of the Real-World Progression in Diabetes (RAPIDS) Model. Med Decis Making 2019;39:137–51. DOI: 10.1177/0272989X18817521.

5. Hayes AJ, Leal J, Gray AM, Holman RR, and Clarke PM. UKPDS outcomes model 2: a new version of a model to simulate lifetime health outcomes of patients with type 2 diabetes mellitus using data from the 30 year United Kingdom Prospective Diabetes Study: UKPDS 82. Diabetologia 2013;56:1925–33. DOI: 10.1007/s00125-013-2940-y.

6. Pandya A, Sy S, Cho S, Alam S, Weinstein MC, and Gaziano TA. Validation of a Cardiovascular Disease Policy Microsimulation Model Using Both Survival and Receiver Operating Characteristic Curves. Med Decis Making 2017;37:802–14. DOI: 10.1177/0272989X17706081.

7. Rizopoulos D. Joint Models for Longitudinal and Time-to-Event Data: With Applications in R. New York: Chapman and Hall/CRC Press, 2012.

8. Freedberg KA, Scharfstein JA, Seage G. R. 3, et al. The cost-effectiveness of preventing AIDS-related opportunistic infections. JAMA 1998;279:130–6. DOI: 10.1001/jama.279.2.130.

9. Pandya A, Sy S, Cho S, Weinstein MC, and Gaziano TA. Cost-effectiveness of 10-Year Risk Thresholds for Initiation of Statin Therapy for Primary Prevention of Cardiovascular Disease. JAMA 2015;314:142–50. DOI: 10.1001/jama.2015.6822.

10. Anderson KM, Wilson PW, Odell PM, and Kannel WB. An updated coronary risk profile. A statement for health professionals. Circulation 1991;83:356–62. DOI: 10.1161/01.cir.83.1.356.

11. Goff DC, Lloyd-Jones DM, Bennett G, et al. 2013 ACC/AHA Guideline on the Assessment of Cardiovascular Risk. Circulation 2014;129. Publisher: American Heart Association:S49–S73. DOI: 10.1161/01.cir.0000437741.48606.98. URL: 10.1161/01.cir.0000437741.48606.98 (visited on 10/28/2023).

12. The Atherosclerosis Risk in Communities (ARIC) Study: design and objectives. The ARIC investigators. Am J Epidemiol 1989;129:687–702.

13. Bild DE, Bluemke DA, Burke GL, et al. Multi-Ethnic Study of Atherosclerosis: objectives and design. Am J Epidemiol 2002;156:871–81. DOI: 10.1093/aje/kwf113.

14. Leal J, Alva M, Gregory V, et al. Estimating risk factor progression equations for the UKPDS Outcomes Model 2 (UKPDS 90). Diabet Med 2021;38:e14656. DOI: 10.1111/dme.14656.

15. Bates D, Mächler M, Bolker B, and Walker S. Fitting Linear Mixed-Effects Models Using lme4. Journal of Statistical Software 2015;67:1–48. DOI: 10.18637/jss.v067.i01.

16. Rizopoulos D, Papageorgiou G, and Miranda Afonso P. JMbayes2: Extended Joint Models for Longitudinal and Time-to-Event Data. 2023. URL: https://drizopoulos.github.io/JMbayes2/.

17. Fauvernier M, Remontet L, Uhry Z, Bossard N, and Roche L. survPen: an R package for hazard and excess hazard modelling with multidimensional penalized splines. Journal of Open Source Software 2019;4:1434. DOI: 10.21105/joss.01434.

18. Austin PC, Harrell F. E. J, and Klaveren D van. Graphical calibration curves and the integrated calibration index (ICI) for survival models. Stat Med 2020;39:2714–42. DOI: 10.1002/sim.8570.

19. Austin PC, Putter H, Giardiello D, and Klaveren D van. Graphical calibration curves and the integrated calibration index (ICI) for competing risk models. Diagn Progn Res 2022;6:2. DOI: 10.1186/s41512-021-00114-6.

20. R Core Team. R: A Language and Environment for Statistical Computing. R Foundation for Statistical Computing. Vienna, Austria, 2021. URL: https://www.R-project.org/.

21. Siebert U, Alagoz O, Bayoumi AM, et al. State-transition modeling: a report of the ISPOR-SMDM Modeling Good Research Practices Task Force-3. Med Decis Making 2012;32:690–700. DOI: 10.1177/0272989X12455463.

22. Lau B, Cole SR, and Gange SJ. Competing risk regression models for epidemiologic data. Am J Epidemiol 2009;170:244–56. DOI: 10.1093/aje/kwp107.

23. Sadatsafavi M, Yoon Lee T, and Gustafson P. Uncertainty and the Value of Information in Risk Prediction Modeling. Med Decis Making 2022;42:661–71. DOI: 10.1177/0272989X2-21078789.

24. Menzies NA, Soeteman DI, Pandya A, and Kim JJ. Bayesian Methods for Calibrating Health Policy Models: A Tutorial. Pharmacoeconomics 2017;35:613–24. DOI: 10.1007/s40273-017-0494-4.

## References

1. The Atherosclerosis Risk in Communities (ARIC) Study: design and objectives. The ARIC investigators. Am J Epidemiol 1989;129:687–702.

2. Wright JD, Folsom AR, Coresh J, et al. The ARIC (Atherosclerosis Risk In Communities) Study: JACC Focus Seminar 3/8. J Am Coll Cardiol 2021;77:2939–59. DOI: 10.1016/j.jacc.2021.04.035.

3. Bild DE, Bluemke DA, Burke GL, et al. Multi-Ethnic Study of Atherosclerosis: objectives and design. Am J Epidemiol 2002;156:871–81. DOI: 10.1093/aje/kwf113.

4. MESA Study Timeline and Procedures. 2023. URL: https://www.mesa-nhlbi.org/aboutMESAStudyTime.aspx (visited on 03/21/2023).

5. Rizopoulos D, Papageorgiou G, and Miranda Afonso P. JMbayes2: Extended Joint Models for Longitudinal and Time-to-Event Data. 2023. URL: https://drizopoulos.github.io/JMbayes2/.

6. Rizopoulos D. Joint Models for Longitudinal and Time-to-Event Data: With Applications in R. New York: Chapman and Hall/CRC Press, 2012.

7. Rizopoulos D. The R Package JMbayes for Fitting Joint Models for Longitudinal and Time-to-Event Data Using MCMC. Journal of Statistical Software 2016;72:1–46. DOI: 10.18637/jss.v072.i07. URL: https://www.jstatsoft.org/index.php/jss/article/view/v072i07.

8. Fauvernier M, Remontet L, Uhry Z, Bossard N, and Roche L. survPen: an R package for hazard and excess hazard modelling with multidimensional penalized splines. Journal of Open Source Software 2019;4:1434. DOI: 10.21105/joss.01434.

9. Fauvernier M, Roche L, Uhry Z, et al. Multi-Dimensional Penalized Hazard Model with Continuous Co-variates: Applications for Studying Trends and Social Inequalities in Cancer Survival. Journal of the Royal Statistical Society Series C: Applied Statistics 2019;68:1233–57. DOI: 10.1111/rssc.12368.

10. Leal J, Alva M, Gregory V, et al. Estimating risk factor progression equations for the UKPDS Outcomes Model 2 (UKPDS 90). Diabet Med 2021;38:e14656. DOI: 10.1111/dme.14656.

11. Pandya A, Sy S, Cho S, Alam S, Weinstein MC, and Gaziano TA. Validation of a Cardiovascular Disease Policy Microsimulation Model Using Both Survival and Receiver Operating Characteristic Curves. Med Decis Making 2017;37:802–14. DOI: 10.1177/0272989X17706081.

12. Andrinopoulou ER, Rizopoulos D, Takkenberg JJM, and Lesaffre E. Combined dynamic predictions using joint models of two longitudinal outcomes and competing risk data. Statistical Methods in Medical Research. 26:1787–801. DOI: 10.1177/0962280215588340.

13. Austin PC, Putter H, Giardiello D, and Klaveren D van. Graphical calibration curves and the integrated calibration index (ICI) for competing risk models. Diagn Progn Res 2022;6:2. DOI: 10.1186/s41512-021-00114-6.

